# The characteristics of multi-source mobility datasets and how they reveal the luxury nature of social distancing in the U.S. during the COVID-19 pandemic

**DOI:** 10.1101/2020.07.31.20143016

**Authors:** Xiao Huang, Zhenlong Li, Yuqin Jiang, Xinyue Ye, Chengbin Deng, Jiajia Zhang, Xiaoming Li

**Affiliations:** Department of Geosciences, University of Arkansas; Geoinformation and Big Data Research Laboratory, Department of Geography, University of South Carolina; Department of Landscape Architecture & Urban Planning, Texas A&M University; Department of Geography, State University of New York at Binghamton; Epidemiology and Biostatistics, Arnold School of Public Health, University of South Carolina; Health Promotion, Education, and Behavior, Arnold School of Public Health, University of South Carolina

**Keywords:** COVID-19, Mobility, Responsive index, Visualization

## Abstract

This study reveals the human mobility from various sources and the luxury nature of social distancing in the U.S during the COVID-19 pandemic by highlighting the disparities in mobility dynamics from lower-income and upper-income counties. We collect, process, and compute mobility data from four sources: 1) Apple mobility trend reports, 2) Google community mobility reports, 3) mobility data from Descartes Labs, and 4) Twitter mobility calculated via weighted distance. We further design a Responsive Index (*RI*) based on the time series of mobility change percentages to quantify the general degree of mobility-based responsiveness to COVID-19 at the U.S. county level. We find statistically significant positive correlations in the *RI* between either two data sources, revealing their general similarity, albeit with varying Pearson’s *r* coefficients. Despite the similarity, however, mobility from each source presents unique and even contrasting characteristics, in part demonstrating the multifaceted nature of human mobility. The positive correlation between *RI* and income at the county level is significant in all mobility datasets, suggesting that counties with higher income tend to react more aggressively in terms of reducing more mobility in response to the COVID-19 pandemic. Most states present a positive difference in *RI* between their upper-income and lower-income counties, where diverging patterns in time series of mobility changes percentages can be found. To our best knowledge, this is the first study that cross-compares multi-source mobility datasets. The findings shed light on not only the characteristics of multi-source mobility data but also the mobility patterns in tandem with the economic disparity.

**Highlights:**

- Human mobility data provide valuable insight into how we adjust our travel behaviors during the COVID-19 pandemic.
- Human mobility records from Descartes Labs, Apple, Google, and Twitter are compared.
- Multi-source mobility datasets well capture the general impact of COVID-19 pandemic on mobility in the U.S. but present unique and even contrasting characteristics
- The proposed responsive index quantifies the level of mobility-based reaction in response to the COVID-19 pandemic
- All selected mobility datasets suggest a statistically significant positive correlation between the responsive index and median income at the U.S. county level.

## 1. Introduction

The outbreak of Coronavirus disease (COVID-19) has undoubtedly raised worldwide concerns. On March 11, the World Health Organization (WHO) officially declared COVID-19 as a pandemic, pointing to the sustained risk of further global spread (WHO, 2020a). As the new epicenter of the COVID-19, there had been 4,405,932 cases and 150,283 deaths in the U.S by July 30, 2020, according to the Centers for Disease Control and Prevention (CDC). They accounted for 26.2% and 22.7% of the global statistics (WHO, 2020b). As the COVID-19 pandemic progresses, social distancing, one of the non-pharmacological control measures to reduce person-to-person contact, has emerged as an effective measure to restrain the spread of infections (Jawaid, 2020). Studies discovered that the considerable mobility reduction following the implementation of social distancing measures is greatly responsible for the reduction of the transmission of SARS-COV-2, the virus that caused the COVID-19 pandemic (Kraemer et al., 2020; Qiu et al., 2020; Buckee et al., 2020; Oliver et al., 2020). However, the disproportionate responses in mobility due to the different socioeconomic status reflects the long-standing disparities in health outcomes and potentially leave more vulnerable populations uniquely exposed to the COVID-19 pandemic (Almagro and Orane-Hutchinson, 2020; Coven and Gupta, 2020).

The investigation of inequality and disease is not new in literature, as many pieces of epidemiological evidence that prove a robust relationship between social inequality and health outcomes have been found (Nguyen and Peschard, 2003; Muennig et al., 2005; Cooper, 2001; Gaziano et al., 2010). Income, particularly, as one of the major factors in socioeconomic status, is responsible for the noticeable disparities in the exposure of many diseases (Siddharthan et al., 2018; Muennig et al., 2005; Breteler et al., 2013). For the COVID-19 pandemic, Barnett-Howell and Mobarak (2020) found that the epidemiological and economic benefits of social distancing are much smaller in poorer countries, as the poor place relatively greater value on their livelihood concerns compared to contracting COVID-19. A recent study by Nayak et al. (2020) links the socioeconomic vulnerability with mortality rates in U.S. counties. They find that counties with higher social vulnerability (primarily driven by low income) are experiencing greater mortality rates. By investigating the role of income inequality in moderating the effectiveness of social distancing measures, Chiou and Tucker (2020) find that high-income earners are more likely to self-isolate at home, and the evidence further suggests that the presence of high-speed Internet plays an important role. What makes things worse is that the existing income-induced disparities in the responses of COVID-19 tend to be exaggerated by the recognized shortcomings of the U.S. protection measures (e.g., health insurance, minimum incomes, unemployment benefits, and paid parental leave), potentially causing long-term negative outcomes for the low-income populations (Coven and Gupta, 2020; Lou et al., 2020). Despite the above efforts, multi-source and multiscale evidence is still needed to understand whether/how the wealthy and the poor populations respond to the COVID-19 pandemic differently.

Existing research has suggested that the disproportionate exposures to the risk from the COVID-19 between the lower-income and upper-income groups can be properly measured using the human mobility data, given the close relationship between human mobility and the transmission of SARS-COV-2 (Tian et al., 2020; Xu and Li, 2020; Fauver et al., 2020). Since the outbreak of COVID-19, many mobility data sources have been made available to facilitate rapid monitoring in human mobility, most notably Google mobility report derived from Google Maps (www.google.com/covid19/mobility) and Apple mobility report derived from Apple Maps (www.apple.com/covid19/mobility). In addition to the mobility data collected from cellphone navigation applications, mobile network operators start to collaborate with local authorities and the federal government to estimate the impact of mobility-reducing related measures (Oliver et al., 2020; Scott et al., 2020). One notable effort is by Descartes Labs (www.descarteslabs.com), a platform that has open-sourced the daily mobility statistics in the U.S. collected via mobile devices (Warren and Skillman, 2020). As a more harmonized and less privacy-concerning data source, social media data (e.g., Twitter and Facebook) are also favored by many scholars to study the mobility dynamics during the COVID-19 pandemic (e.g., Huang et al., 2020; Yang et al., 2020). Given the existence of publicly available mobility datasets from various sources, understanding their similarities and dissimilarities is in great need. Owing to the multifaceted nature of human mobility (Gonzalez et al., 2008), however, neither cellular records, navigation applications, nor social media, can solely represent human mobility as a whole. Instead, they reflect human mobility from varying yet valuable perspectives. Linking these perspectives with economic disparities contributes to a better understanding of the mobility dynamics of groups with different socioeconomic status in response to the COVID-19 pandemic.

In this study, we aim to examine 1) the similarity and dissimilarity of mobility from various sources, and 2) the luxury nature of social distancing in the U.S during the COVID-19 pandemic by highlighting the disparities in mobility dynamics from lower-income and upper-income groups. We collect and compute mobility data from a variety of sources, including Google and Apple mobility reports (navigation applications), Descartes Labs mobility (cellular records), and Twitter (social media). To quantify the general degree of mobility changes at the county level, we designed a responsive index (*RI*) via the time series of mobility change percentage using the sources above. Specifically, we attempt to answer whether a consensus can be reached from various mobility sources that the lower- and upper-income groups present contrasting mobility dynamics during the pandemic, eventually leading to disproportionate exposures that disfavor the lower-income group. Our study extends the increasing amount of literature in understanding disparities via big mobility data. The findings of this study help us gain knowledge of not only the similarities and dissimilarities in multi-source mobility data but also the wealth disparity in tandem with the implementation of social distancing, greatly benefiting epidemic modeling and policy-making for better mitigation of future pandemics.

## 2. Mobility datasets and preprosessing

We collect and compute four open-source mobility datasets that cover the U.S., which include 1) Mobility records from Descartes Labs using commercially available mobile device dataset; 2) Apple mobility reports mainly from Apple Maps; 3) Google community mobility reports mainly from Google Maps, and 4) Twitter-based mobility from geotagged tweets. The finest spatial level of the mobility data from Descartes Labs, Apple, and Google is the U.S. county level. Given that the U.S. National Emergency was announced on March 13 and the majority of the U.S. states started to react aggressively after mid-March, we present the daily mobility change percentage from March 1, 2020, to June 30, 2020. We believe that this four-month period well covers different epidemic phases in the U.S., thus providing valuable knowledge of how people react to the COVID-19 pandemic by adjusting their travel behaviors accordingly. Although the four mobility datasets in this study differ from each other in terms of data quantity, data quality, and baseline calculations, we apply several preprocessing steps to make them more comparable (as detailed in the following subsections). To ensure that data records are sufficient enough to generate reliable and stable time series of mobility changes, we only map the time series for counties with more than 100 days of mobility records (out of 122 days from March 1 to June 30). To fill the missing data, we apply a simple linear interpolation, under the assumption that mobility changes linearly between two consecutive available records.

### 2.1 Mobility data from Descartes Labs

The mobility dataset from Descartes Labs, a predictive intelligence company that makes data-agnostic platforms for large-scale analysis, is open-sourced at GitHub (https://github.com/descarteslabs/DL-COVID-19) and updated daily. The data cover a total of 2,668 counties. Among them, 2,612 counties have mobility records in more than 100 days from March 1 to June 30). The data are derived from a collection of mobile devices reporting consistently throughout the day. The distance measured in this dataset is the daily maximum distance of a certain user, i.e., the maximum distance between a user’s initial location of a day and other locations within the same day (details can be found in Warren and Skillman et al. (2020)). The county-level mobility baseline is defined as the median of the maximum distance of all users in a certain county on weekdays from February 17, 2020, to March 7, 2020. The mobility change percentage is further calculated by comparing daily mobility to the baseline. Due to the data quality issues, mobility data on April 20 and May 29 are not released (Warren and Skillman et al., 2020). To facilitate a smooth time series mapping, we generate mobility change percentage on these two missing days for all available counties by averaging the values of the corresponding days in the preceding week and following week. That is, the mobility change percentage for April 20 is the average value of mobility change percentages on April 13 and April 27, and May 29 is the average of May 22 and June 5.

### 2.2 Apple mobility reports

The raw mobility reports from Apple (www.apple.com/covid19/mobility) cover major cities and a total of 63 countries (the U.S. included) and regions. Unlike Descartes Labs’ mobility data that measure the travel distance, Apple mobility reports are generated by counting the number of requests made to Apple Maps for directions (Apple Mobility Trends Reports, 2020). Despite the difference in measurement, the daily changes in the number of requests from navigation services like Apple Maps still offer valuable insights into people’s mobility changes in response to the COVID-19 pandemic. Although Apple provides mobility records in three different categories that include “transit”, “walking”, and “driving”, only “driving” is available for the U.S. at the county level. Apple mobility reports cover 2,070 U.S. counties, and all the counties have mobility records in more than 100 days in the designated period. However, data for May 11 and May 12 are not available. Following the same procedure in the Descarte Labs mobility, we generate mobility data of the two missing days by averaging the corresponding days in the preceding week and the following week. In addition, Apple defines the mobility from January 13 as the baseline value. To make it comparable with Descartes Labs mobility, we compute a new baseline value from February 17 to March 7 (the baseline period defined in Descartes Labs mobility) and use it to adjust the mobility in the entire Apple mobility dataset.

### 2.3 Google mobility reports

Google mobility reports use aggregated, anonymized data to chart movement trends over time, across six different categories in “retail and recreation”, “groceries and pharmacies”, “parks”, “transit stations”, “workplaces”, and “residential” (Google Community Mobility Reports, 2020). Although a total of 2,794 U.S. counties are covered, our investigation reveals that the quantity of the records varies greatly across categories, with “workplaces” having the most records and other categories insufficient for deriving stable time series. Therefore, we select mobility in the category of “workplaces” for further analysis. Google defines the baseline of the dataset as the median value for the corresponding day of the week, during the 5-week period January 3 to February 6. To make it comparable with other datasets, we adjust the Google mobility records by computing a new baseline value from February 17 to March 7. Within 2,794 counties covered by the dataset, 2,110 counties have records in more than 100 days from March 1 to June 30 and are therefore selected to derive the mobility time series.

### 2.4 Twitter-based mobility

We have collected about 197 million geotagged (embedded with geolocation in the format of exact coordinates or place names) tweets in a time period from January 1, 2020, to June 30, 2020, from over 2.9 million unique Twitter users in the U.S. using the official Twitter Streaming Application Programming Interface (API). Following the work by Huang et al. (2020), we compute two types of distances 1) single-day distance (*d*_*s*_) that reveals daily travel patterns and 2) cross-day displacement (*d*_*c*_) that reveals users’ displacement between two consecutive days. We then calculate a weighted distance(*d*_*w*_) of each county by integrating these two distances (Equation 1).

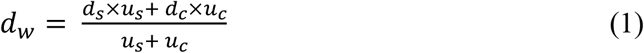

where *u*_*s*_ and *u*_*c*_ denote the user counts for the calculation of *d*_*s*_ and *d*_*c*_ in each county, respectively. By considering both single-day and cross-day distance, the weighted distance is expected to reflect the overall mobility dynamics from Twitter. Our investigation suggests that a stable time series can be achieved when the daily user count for the distance calculation reaches 30 (see Figure A in the Appendices). Within the 2,981 counties covered by the Twitter dataset, 565 counties are qualified for the time series mapping as their daily user counts are greater than or equal to 30 for more than 100 days between March 1 and June 30. Following the baseline settings of the aforementioned mobility datasets, we compute the mobility change percentage by setting the baseline between February 17 to March 7.

## 3. Methods

### 3.1 Conceptualization of responsive index (RI)

The time series of the mobility change percentage from the various mobility sources generally quantify the level of reaction in response to the COVID-19 pandemic. A reduction in mobility denotes the positive response while an increase in mobility denotes otherwise. The strength of the response is assumed to be proportional to the degree in mobility changes. We use baseline mobility to divide the responsiveness into positive and negative space. Different from the mobility-based responsiveness in Huang et al. (2020), we calculate the ratios respectively in positive response and negative response to confine our *RI* into a range of [-1,1]. Figure 1 describes the concept of the responsive index (*RI*) using time series in mobility change percentage.

**Figure 1.**
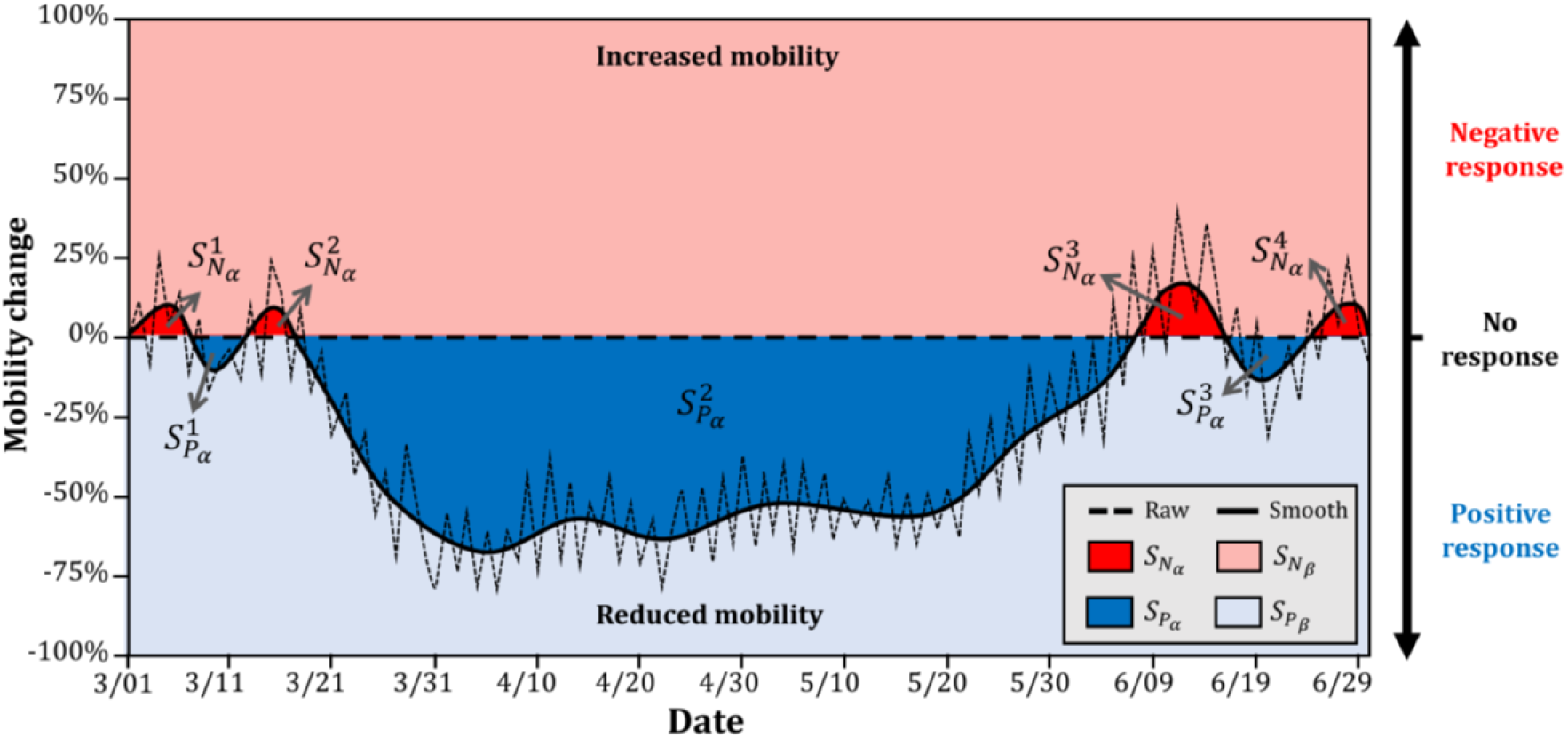
The concept of the responsive index using time series in mobility changes.

We first smooth the time series using a one-dimensional Gaussian filter to remove the noises so that the general trend of mobility changes is revealed. The baseline (representing no mobility change) divides the space into two parts: positive response with reduced mobility (*S*_*P*_) and negative response with increased mobility (*S*_*P*_) (Figure 1). The area between the baseline and the smooth time series is denoted as *α*, while space excepts *α* is denoted as *β*. In Figure 1, there are three individual 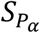 regions, respectively denoted as 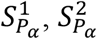 and 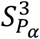, and four 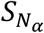 regions, respectively denoted as 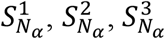, and 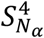. We then quantify the response by computing two ratios: 1) 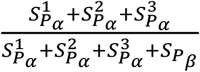 and 2) 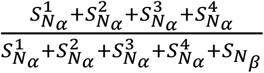, where the former ratio quantifies the strength of the positive response, and the latter ratio quantifies the strength of the negative response (Figure 1). The *RI* we propose is the difference between the two ratios. Given that Figure 1, for the illustration purpose, confines the mobility change in a range from -100% to 100%, we have 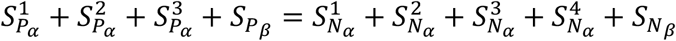. In this study, however, the mobility change percentage is unrestricted from 0% to 100% in the calculation of *RI*. Generally, the *RI* can be calculated via the following equation:

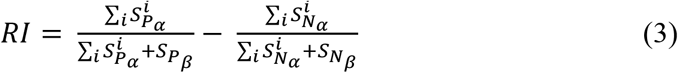

Our proposed *RI* has a range of [-1, 1] with *RI* = 1 suggesting a hypothetically perfect responsiveness. Positive *RI* suggests that the accumulative response within a specific period of time is positive, while negative *RI* suggests otherwise. The higher the *RI*, the stronger the accumulative responsiveness a geographic region has.

### 3.2 Comparison of mobility datasets and their association with income

In this study, we aim to cross-compare mobility datasets from various sources and reveal the linkage between mobility dynamics and income at the U.S. county level, as a county is the finest geographical unit in mobility datasets from Apple, Google, and Descartes. Let *MCP*_*j*_ denote the mobility change percentage of a certain county on day *j*. We derive the heat map by plotting all the available pairs, i.e., 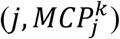, within the time frame (March 1 to June 30) for each mobility dataset. In addition, we apply the Pearson correlation analysis for the four open-source mobility datasets to determine 1) the correlation of the derived county-level *RI* from each pair of mobility sources, and 2) the correlation between county median income (from the latest ACS 5-year (2014-2018) estimates) and the county-level *RI*. To reveal the disparity in mobility changes between lower-income and upper-income counties, we first rank the available counties in each dataset based on the median income and derive the time series of mobility change percentages respectively for lower-income and higher-income counties by selecting the median values. Given the fact that state policies may greatly shape county-level mitigation strategies, we also visualize the statewide disparity in the time series of mobility change percentages and the difference in *RI* for upper-income counties and lower-income counties within each CONUS state to remove the potential impact resulting from the statewide policies.

## 4. Results

### 4.1 Comparison of the four mobility datasets

In general, the impact of the COVID-19 pandemic is well documented from all four mobility datasets, evidenced by the clear deviations in mobility from the baseline since mid-March (Figure 2), when the WHO declared COVID-19 as a pandemic (March 11) and the United States declared a National Emergency (March 13). In April, mobility from all four datasets descended to the bottom and remained considerably lower than baseline throughout the month. With the gradually loosened measures from the “Opening Up America Again” guidelines, the mobility gained an upward momentum and started to bounce back in early-May, resulting in the “U” shape distribution that can be observed from all four datasets. Despite the similarity in the general trend, mobility from each source presents unique and even contrasting characteristics. Data from Apple mobility trend reports show that mobility in the U.S. had returned to the baseline in mid-May and remained above the baseline in the entire June (Figure 2a), which contradicts the mobility dynamics from the other three datasets where mobility is found not fully recovered to the pre-pandemic level in May and June (Figure 2b-d). This characteristic of the Apple mobility dataset can be partially explained by its single-day (January 13) baseline setting. Although we adjust Apple’s baseline by extending the baseline temporal coverage, the intrinsic monthly discrepancies in mobility between January and other months still exist. That is, the above-the-baseline mobility in May and June is arguably the exaggeration that results from the low baseline value observed on January 13. Compared with March and April, the mobility change percentages show a more dispersed distributing pattern in May and June (Figure 3a), indicating the inconsistency of counties in the recovering phase. The dataset from Descartes Labs also suggests a strong recovery in mobility following the lifting of strict measures since early-May (Figure 2b). In comparison to the other three datasets, the reduction is more obvious in April, evidenced by the high concentration close to -100% in mobility change. We investigate the counties with barely any recorded mobility in April and find that they coincide well with the counties that were the epicenter and faced strict stay-at-home orders (e.g., New York, Queens, and Kings in the State of New York, and Alameda and Santa Clara in California). The close-to-zero mobility in these counties can be explained by the data sources (i.e., commercially available mobile device location dataset) (Warren and Skillman, 2020), which own a more passive nature compared to the data collected via smartphone applications. Despite the slight recovery in May and June, Google mobility shows a more concentrated distribution that mostly remains below the baseline throughout the time period, suggesting that the recovery of mobility in the category of “workplaces” is presumably slower than other types. In addition, the reduction in April from Google mobility is relatively less dramatic compared with other datasets. Our Twitter-based mobility dataset reveals a similar reducing-and-recovering pattern (Figure 2d), although its heat map presents a rather scattered distribution largely due to the uncertainties introduced from the limited number of geotagged tweets in certain counties. Despite the scattered distribution pattern, the significant mobility drop in mid-March and the constantly low mobility in April are well captured by the heat map (Figure 2d).

**Figure 2.**
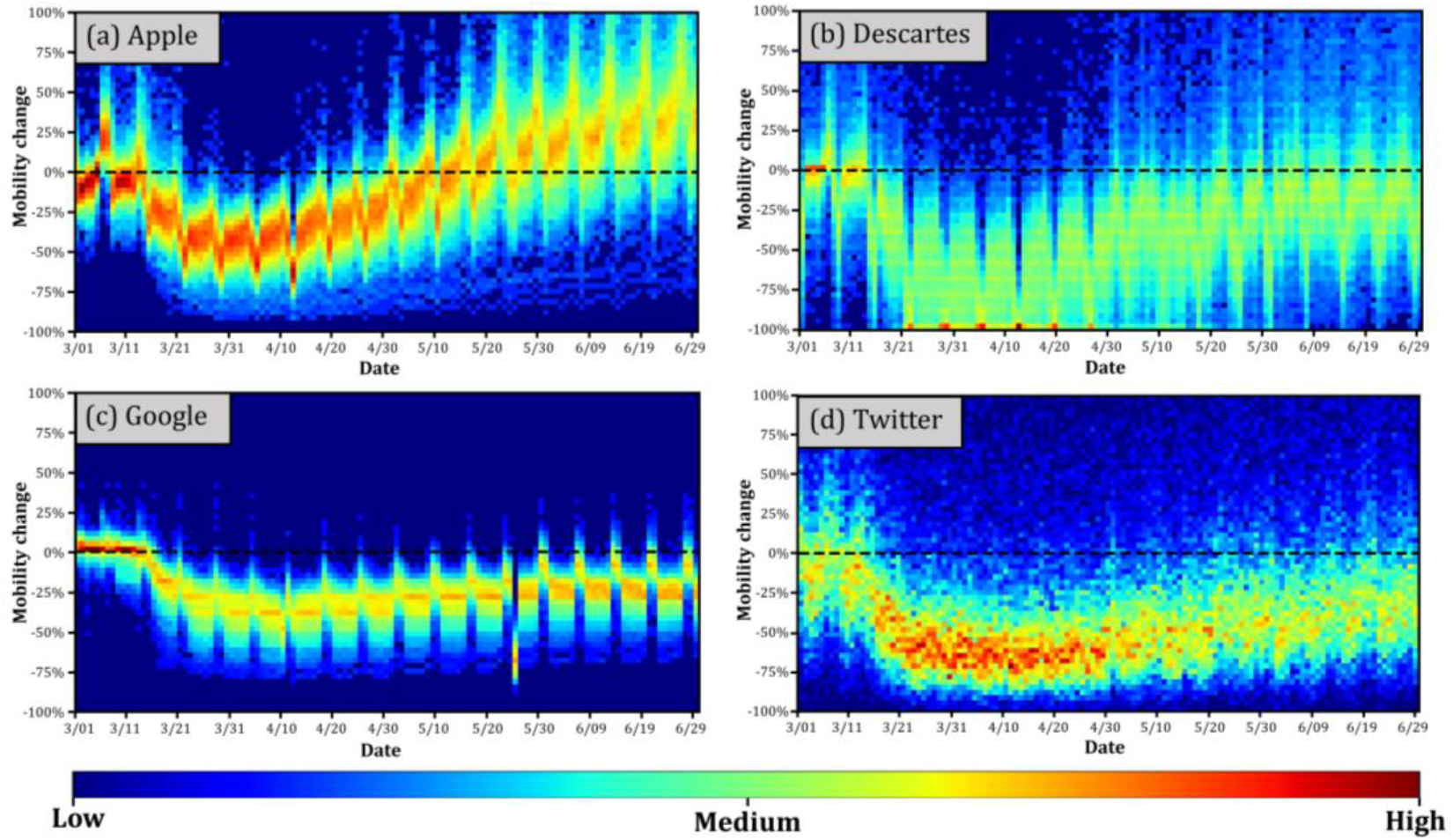
Heat map of the county-level mobility change percentages for four sources (confined from -100% to 100%). (a) Apple mobility trend reports; (b) Mobility data from Descartes Labs; (c) Google community mobility reports; (d) mobility calculated via weighted distance from Twitter.

**Figure 3.**
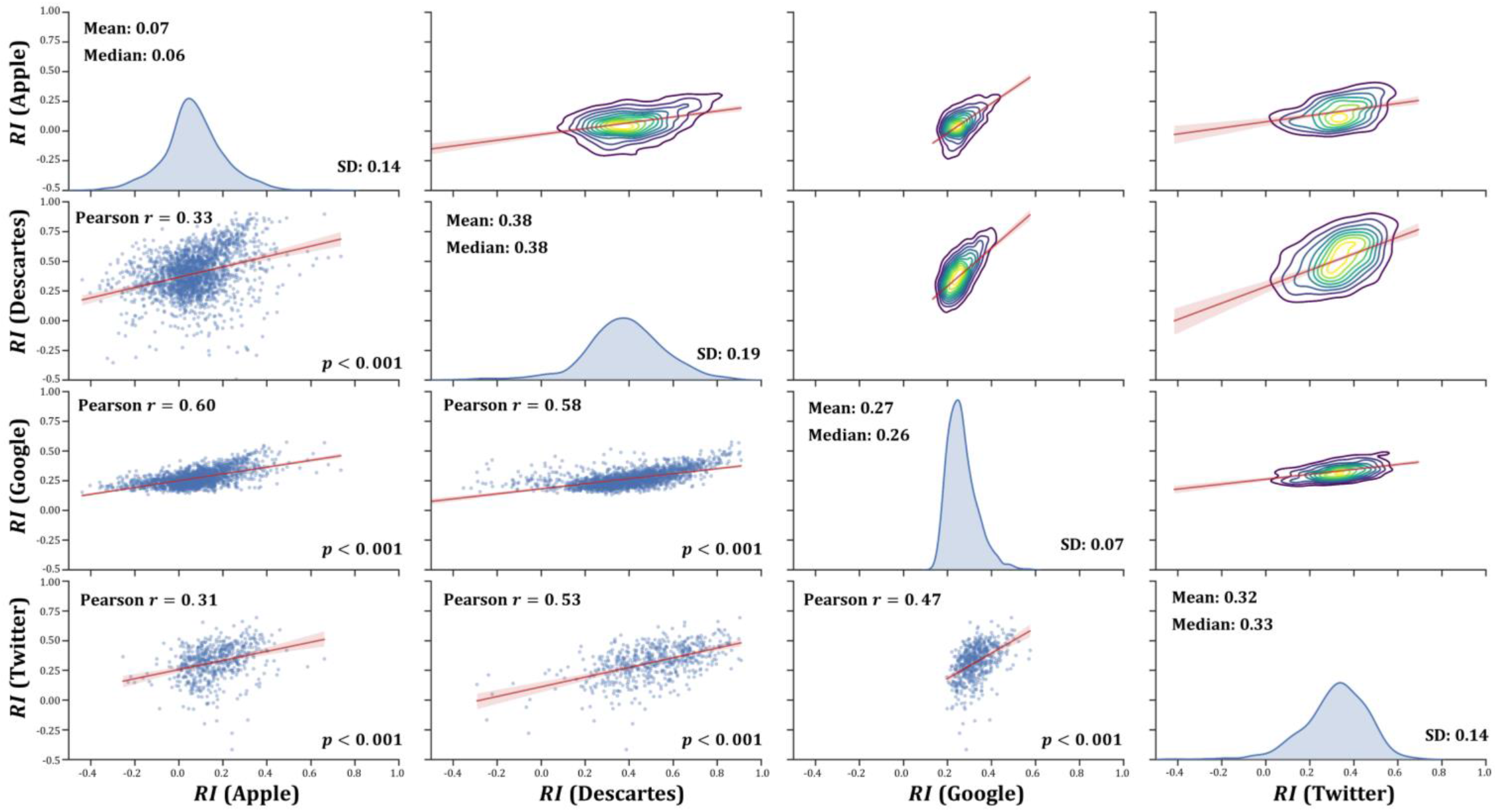
Scatter matrix of county-level Responsive Index (*RI*) calculated from Apple, Descartes, Google, and Twitter mobility datasets. The lower triangle shows the bivariate scatter plots; the upper triangle shows the 2-D kernel density estimations; the diagonal shows the univariate distribution histograms

The results from the Pearson correlation analysis reveal the similarity in computed county-level *RI* from these four mobility datasets (Figure 3). As expected, we find statistically significant positive correlations in *RI* between either two data sources, albeit with varying Pearson’s *r* coefficients (Figure 3). Google and Apple share the highest correlation (*r* = 0.60) owning to the fact that both of their mobility records are from navigation applications in smartphones (i.e., Google Maps and Apple Maps). Despite the high correlation between Apple and Google, however, the correlation between Apple and the other two datasets is relatively weak (*r* = 0.33 for Apple and Descartes, and *r* = 0.31 for Apple and Twitter). Google and Descartes share the second-highest correlation (*r* = 0.58), followed by Twitter and Descartes (*r* = 0.58), and Twitter and Google (*r* = 0.47). The histograms of the four datasets suggest that the distribution of their county-level *RI* follows the normal distribution, but the descriptive statistics vary markedly (Figure 3). Given the low standard deviation, mobility computed from Google Maps reveals less county-level variance in mobility changes in response to the COVID-19 pandemic, which is supported by its concentrated heat map (Figure 2a). The *RI* calculated from Apple mobility (Mean = 0.07 and Median = 0.06) is considerably lower than the other three sources because of documented strong negative responses (above the baseline) in most counties since early-June (Figure 2a). As noted above, however, Apple’s low baseline value on January 13 potentially shifts the entire time series upwards, consequently responsible for the low *RI*s. Twitter (Mean = 0.32 and Median = 0.33) and Descartes (Mean = 0.38 and Median = 0.38) share a similar distribution (histogram shape), indicating that Twitter data is a good proxy of mobile phone data in capturing human mobility.

### 4.2 Correlation between RI and income

Recognizing that the human mobility-based response to the COVID-19 pandemic includes both negative responses (*RI* < 0) and positive responses (*RI* > 0), we explore the correlation between the full spectrum county-level *RI* and the county median income (Figure 4). In general, all mobility datasets reveal a statistically significant positive correlation between *RI* and income, suggesting that counties with higher income tend to react more aggressively in response to the COVID-19 pandemic by reducing more moving activities compared to the pre-pandemic baseline. Nonetheless, we acknowledge that correlation, no matter strong or weak, does not necessarily imply causation. A detailed discussion regarding this issue can be found in Section 5.2. Despite the statistical significance, the strength of the correlation varies among datasets due to the intrinsic nature of their sources. The *RI* calculated from the Google dataset is strongly correlated with income (*r* = 0.65) (Figure 4c1-5c2). In comparison, Apple mobility derived from Apple Maps, also a navigation service, shows a rather weak correlation between its calculated *RI* and income (*r* = 0.29) (Figure 4a1-a2). Three reasons are arguably behind the Google-Apple difference in the strength of the correlation. First, Google mobility in this study only includes the category of “workplaces” (given the insufficient records from other categories) while Apple mobility considers all types of requests. Second, it is reported that the median income for iPhone users in the U.S. is 40% higher than that of Android users (Greenough, 2014). As a result, Apple Maps users possibly do not cover as much demographic spectrum as Google Maps users. Third, the concepts of mobility changes from Apple and Google differ, as Apple measures the change in the number of requests while Google measures the change in the travel distance. Although both concepts are able to reflect the impact of COVID-19 pandemic on human mobility, the difference in calculation potentially causes the difference in the correlation strength. The correlations between income and *RI* calculated from Descartes mobility and Twitter mobility are both significant, and the correlation coefficients are 0.45 (Figure 4b1-b2) and 0.37 (Figure 4d1-d2), respectively. The existence of many outlying counties in datasets from Descartes and Twitter, i.e., counties with strong negative responsiveness (*RI* < 0) (marked in blue bubbles), likely weakens the correlation.

**Figure 4.**
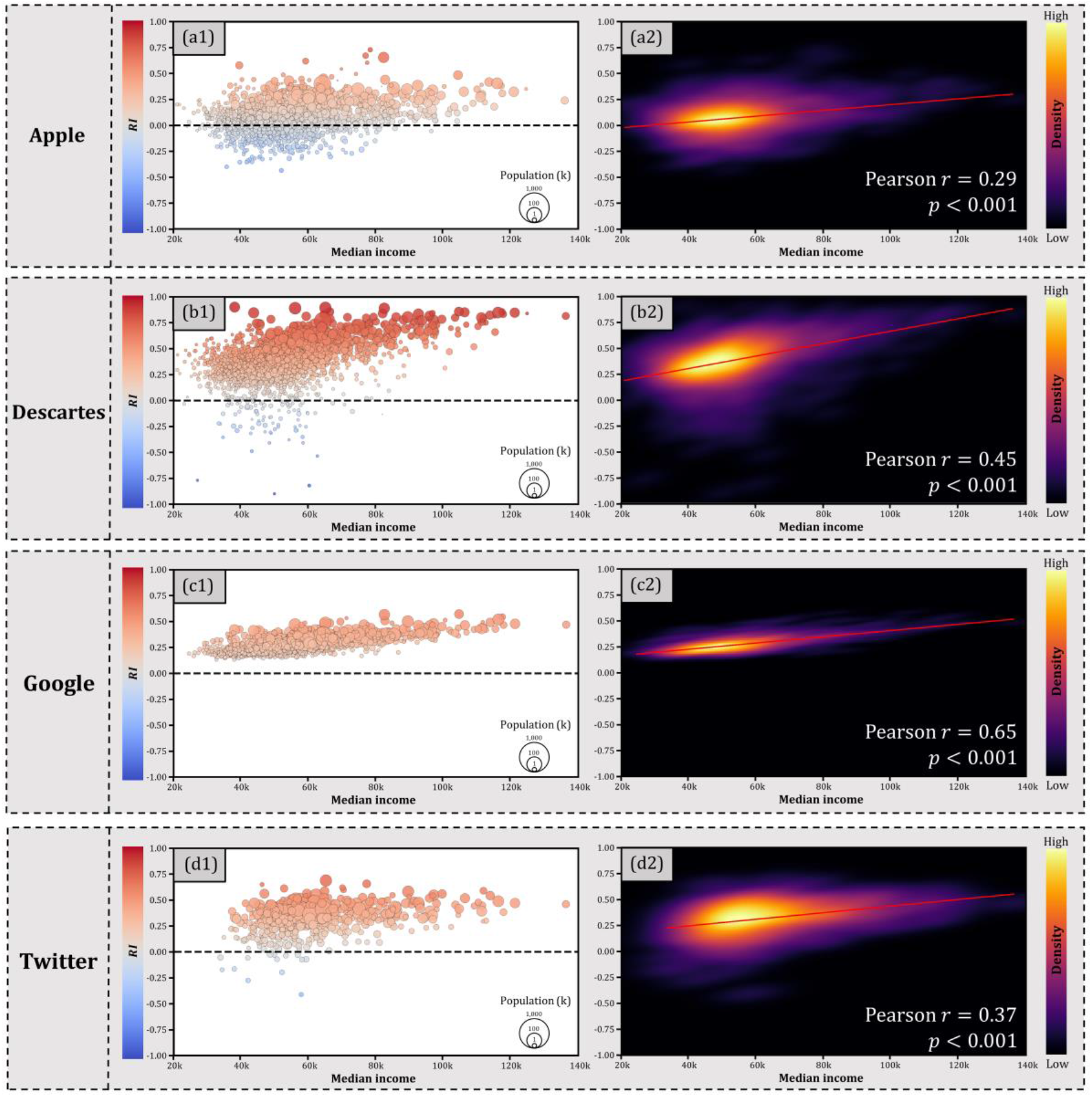
Correlation between RI and income at the county level for four mobility datasets.

### 4.3 Disparities in mobility between lower-income and upper-income counties

We further explore the county-level disparity in mobility dynamics between lower-income and upper-income counties in response to the COVID-19 pandemic. Figure 5 presents the time series of mobility change percentages for all available counties in the four mobility datasets. Specifically, counties in the top 10% and bottom 10% in wealth are highlighted respectively in red lines and blue lines (Figure 5). In general, the disparity in mobility dynamics is well manifested in all mobility datasets, evidenced by the obvious gaps between blue lines and red lines, especially after the declaration of the National Emergency on March 13 (Figure 5). The increase in gaps after March 13 suggests the diverging mobility pattern between lower-income and upper-income counties in response to the pandemic. In addition, rich counties are found more responsive as they present more reduction in mobility compared to poor counties, which is evidenced by the fact that the red line (top 10% in wealth) consistently lies below the blue line (bottom 10% in wealth) in all four mobility datasets. Among the four datasets, Descartes mobility presents the most obvious diverging pattern compared to the other three datasets (Figure 5b). Considering the low standard deviation, however, the separation in Google mobility between the rich and poor counties is also noteworthy, as the red line and blue line are too separated that the gap between them covers most of the time series (Figure 5c). In comparison, the county-level disparities between rich and poor counties from Apple (Figure 5a) and Twitter (Figure 5d) are less dramatic but still noticeable. Note that the number of available counties varies among datasets (e.g., the Twitter dataset contains 565 counties, while the Descartes dataset contains 2,612 counties), which potentially introduces a certain level of inconsistency and uncertainty when heterogeneous mobility data are cross-compared.

**Figure 5.**
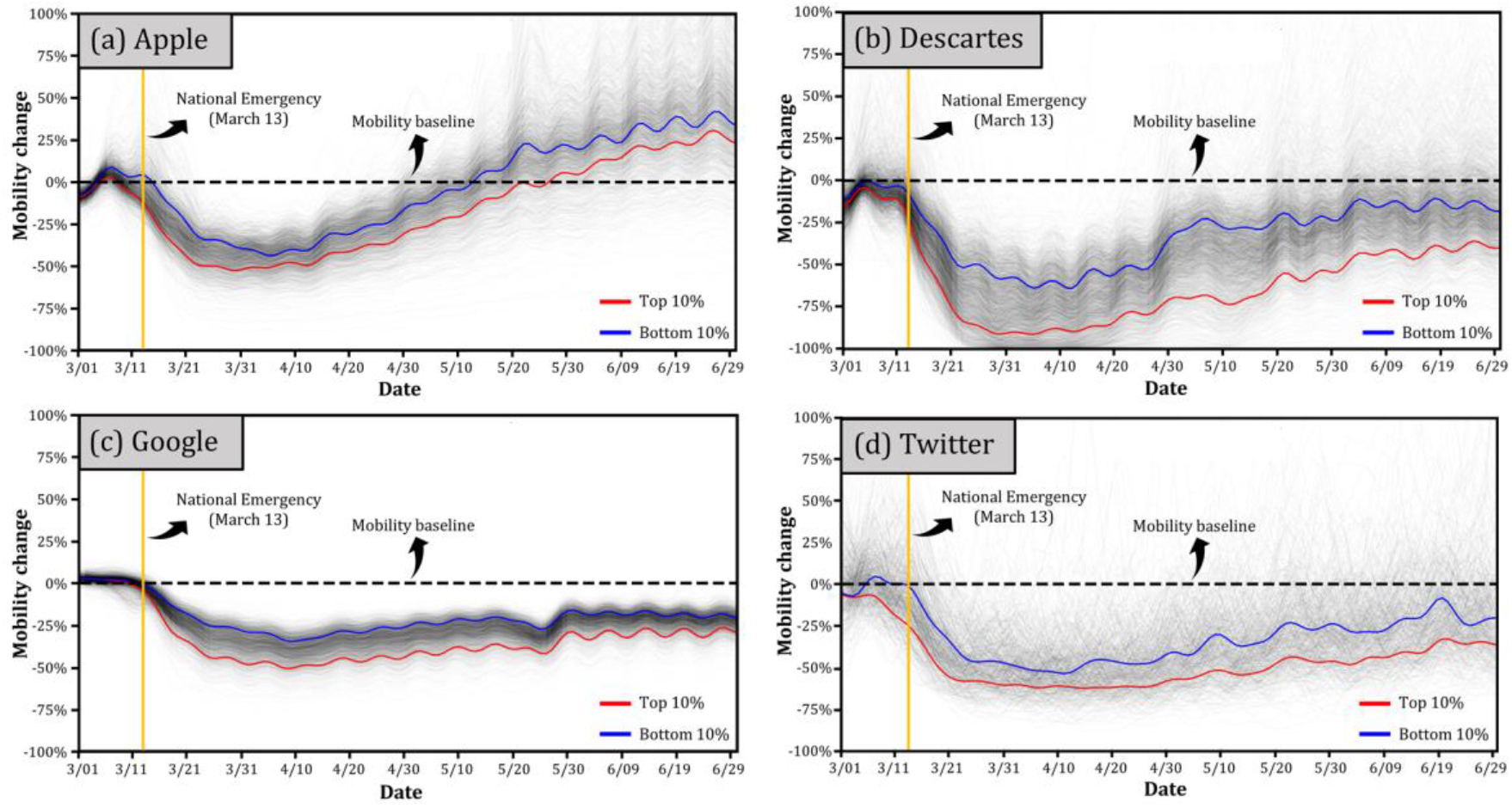
Time series in mobility change percentages of the top 10% and the bottom 10% in wealth for all available counties in the four mobility sources; (a) Apple mobility trend report (2,070 counties); (b) mobility data from Descartes Labs (2,612 counties); (c) Google community reports (2,110 counties); (d) mobility calculated via weighted distance from Twitter (565 counties).

To reveal the disparity in mobility dynamics within each state, we derive the time series of mobility change percentages for the top 20% and bottom 20% counties in wealth within each CONUS state, removing the potential impact from the difference in statewide mitigation policies. The rationale in selecting “20%” (in contrast to the 10% in Figure 5) is to include more counties in each state, especially for states with a small number of counties. Figure 6 shows the statewise time series derived from Descartes mobility in a pseudo-geographical representation of the CONUS states. The results from Apple mobility and Google mobility can be found respectively in Figure A and Figure B in the Appendices. Due to the insufficient number of counties available in each state, the results from Twitter mobility are not presented.

**Figure 6.**
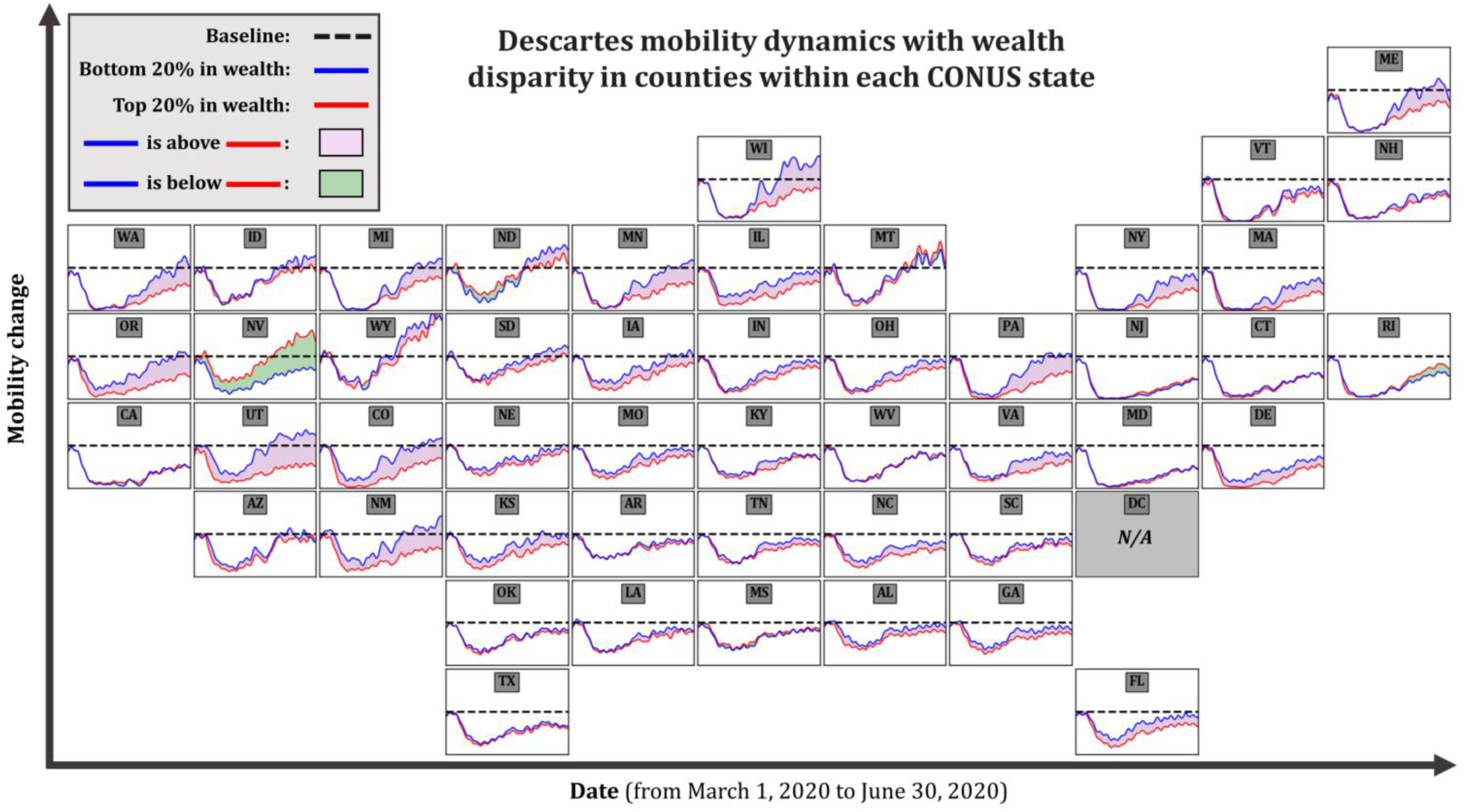
Time series of Descartes mobility change percentages for the top 20% and bottom 20% counties in wealth at each CONUS state (pseudo-geographical representation). For a state with less than five available counties, counties with the most median income and the least median income are selected as the top 20% and the bottom 20%, respectively.

For Descartes mobility, rich counties (the top 20%) within a certain state generally tend to react more aggressively in mobility reduction compared to the poor counties (the bottom 20%) within the same state (Figure 6). The disparity tends to be exaggerated in the mobility recovery phase, i.e., May and June. For example, WA has seen a consistent mobility drop after March 13, suggested by the overlapped blue line and red line. However, the recovery phase in WA highlights the disparity in mobility dynamics, as the poor counties (blue line) obviously gained earlier and greater upward momentum than the rich counties (red line). This similar pattern can also be found in PA, MN, MI, and MA (Figure 6). In comparison, NV stands out by showing the opposite pattern, and states that include LA, TX, CA, MS, and MD present unnoticeable disparity between the rich and the poor counties.

Despite the heterogeneity in sources compared with Descartes mobility, the results from Apple (Figure B) and Google (Figure C) also reveal the general pattern that rich counties tend to be more responsive by showing a higher mobility reduction rate within most CONUS states in the selected time period. However, contrasting patterns, although only a few, can still be found in some states from different mobility data sources. For example, KS presents higher responsiveness for rich counties from Descartes mobility but lower responsiveness for the rich counties from Apple mobility. In addition, the pattern in ND from Google mobility contradicts the pattern from Apple mobility.

To quantitatively reveal the disparity in mobility responsiveness between the rich and the poor counties, we calculate the difference in *RI* between the top 20% and bottom 20% counties in wealth (∇*RI*) at each CONUS state (Figure 7). The values of ∇*RI* are presented in Table 1. In general, states with positive ∇*RI* (in blue) are dominant in number, as 40 states from Descartes mobility, 32 states from Apple mobility, and 42 states from Google mobility, out of the 48 states in the CONUS, are with ∇*RI* > 0 (Figure 8 and Table 1). Mobility datasets from the three sources suggest that, despite the noticeable variances in ∇*RI*, wealthy counties within each state are likely to have more mobility-based responsiveness during the COVID-19 pandemic. However, the inconsistencies from different mobility datasets need to be recognized, as it presumably results from the intrinsic nature of the data sources and therefore warrants caution for further studies that rely on only a single mobility data source.

**Table 1.** The difference in Responsive Index (RI) between the top 20% and bottom 20% counties in wealth at each CONUS state from Descartes mobility, Apple mobility, and Google mobility

| States in the CONUS | $RI_{top\ 20\%} - RI_{bottom\ 20\%}$ measured from | | |
| --- | --- | --- | --- |
|  | Descartes mobility | Apple mobility | Google mobility |
| Alabama | 0.12 | -0.08 | 0.04 |
| Arizona | 0.10 | 0.34 | 0.06 |
| Arkansas | 0.02 | 0.04 | 0.07 |
| California | -0.02 | 0.03 | 0.08 |
| Colorado | 0.32 | 0.39 | 0.17 |
| Connecticut | 0.03 | 0.06 | 0.06 |
| Delaware | 0.18 | 0.09 | 0.07 |
| Florida | 0.18 | -0.02 | 0.09 |
| Georgia | 0.13 | 0.03 | 0.12 |
| Idaho | 0.08 | -0.19 | 0.02 |
| Illinois | 0.22 | 0.03 | 0.08 |
| Indiana | 0.11 | -0.04 | 0.05 |
| Iowa | 0.18 | 0.08 | 0.07 |
| Kansas | 0.19 | -0.09 | 0.04 |
| Kentucky | 0.07 | 0.03 | 0.12 |
| Louisiana | 0.07 | 0.09 | 0.08 |
| Maine | 0.18 | -0.03 | 0.12 |
| Maryland | 0.03 | -0.09 | -0.04 |
| Massachusetts | 0.17 | 0.11 | 0.14 |
| Michigan | 0.15 | 0.08 | 0.11 |
| Minnesota | 0.22 | 0.08 | 0.06 |
| Mississippi | -0.05 | -0.04 | 0.04 |
| Missouri | 0.13 | -0.02 | 0.09 |
| Montana | -0.01 | 0.15 | 0.04 |
| Nebraska | 0.09 | 0.06 | 0.02 |
| Nevada | -0.43 | -0.24 | -0.02 |
| New Hampshire | 0.03 | -0.06 | 0.10 |
| New Jersey | -0.03 | -0.03 | -0.02 |
| New Mexico | 0.34 | 0.20 | 0.11 |
| New York | 0.17 | 0.09 | 0.09 |
| North Carolina | 0.14 | 0.19 | 0.10 |
| North Dakota | -0.01 | 0.06 | -0.13 |
| Ohio | 0.12 | 0.03 | 0.09 |
| Oklahoma | 0.05 | 0.13 | 0.06 |
| Oregon | 0.32 | 0.37 | 0.13 |
| Pennsylvania | 0.27 | 0.13 | 0.11 |
| Rhode Island | -0.05 | -0.15 | -0.03 |
| South Carolina | 0.08 | 0.17 | 0.10 |
| South Dakota | 0.12 | -0.09 | -0.13 |
| Tennessee | 0.09 | 0.13 | 0.09 |
| Texas | 0.05 | 0.05 | 0.07 |
| Utah | 0.45 | 0.06 | 0.01 |
| Vermont | 0.08 | -0.07 | 0.06 |
| Virginia | 0.16 | -0.03 | 0.06 |
| Washington | 0.19 | 0.23 | 0.10 |
| West Virginia | -0.02 | 0.03 | 0.05 |
| Wisconsin | 0.28 | 0.16 | 0.05 |
| Wyoming | 0.11 | 0.01 | 0.01 |

**Figure 7.**
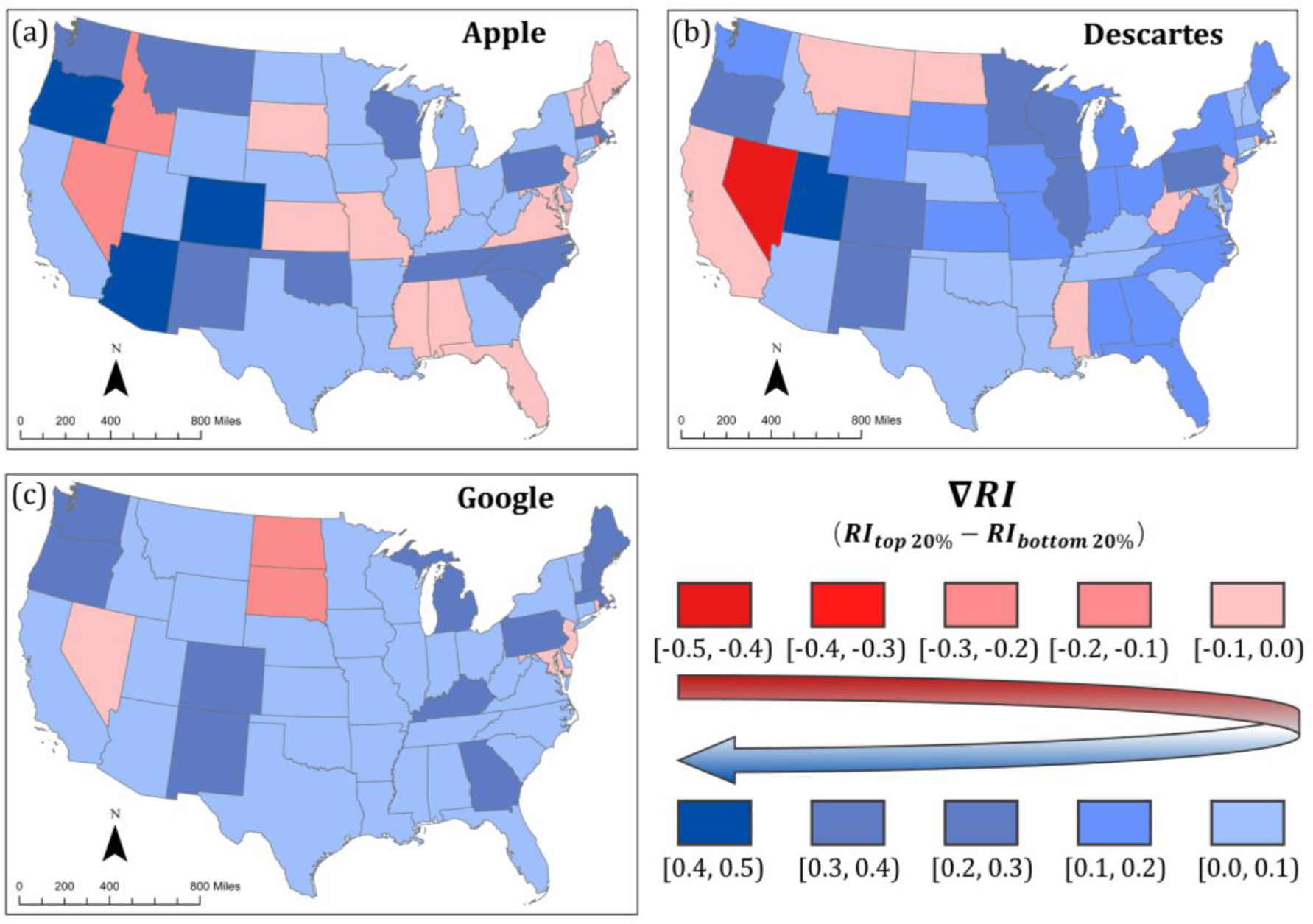
The difference in Responsive Index (RI) between the top 20% and bottom 20% counties in wealth at each CONUS state; (a) Apple mobility trend report; (b) mobility data from Descartes Labs; (c) Google community reports

## 5. Discussions

### 5.1 The fusion value in heterogeneous mobility data

Our investigation in Descartes, Apple, Google, and Twitter mobility reveals that they well capture the impact of the COVID-19 pandemic on human mobility by showing clear deviations from the pre-pandemic baseline. Nevertheless, we observe considerable dissimilarities among the datasets as each source presents unique characteristics. The four selected mobility datasets in this study demonstrate the multifaceted nature of human mobility that has been documented by many (Gonzalez et al., 2008; Cui et al., 2018), and some believe these heterogeneous data sources reflect human mobility from different yet valuable perspectives (Zhang et al., 2014; Lau et al., 2019). Zhang et al. (2014) argue that most of the state-of-the-art theory and practice on human mobility focus on single-source data in isolation from one another, inevitably leading to limited representativeness. In our study, although dataset from Google and Apple are both derived mainly from cellphone navigation applications, the time series in mobility change percentages and the correlation analysis greatly highlight their difference. Since Google Maps is mainly targeted by Android users while Apple Maps by IOS (iPhone Operating System) users, the difference between Google mobility and Apple mobility can be partly explained by the difference in their user’s demographics. Twitter mobility also shows a unique pattern in response to the COVID-19 pandemic, different from all other three mobility datasets. However, we need to acknowledge the bias in Twitter data towards specific groups of the population (Jiang et al., 2019, Martín et al., 2019; Huang et al., 2018).

It is reasonable to assume that the fusion of these mobility datasets mitigates the biases to some degree and provides a holistic view of mobility dynamics in a broad spectrum of the population. Numerous studies have been conducted, attempting to fuse multi-source mobility data towards a more comprehensive one. To list but a few, Zhang et al. (2014) proposed a systematic framework to integrate transit records and cellphone records to mitigate biased sampling. Similarly, Montero et al. (2019) found that the fusion of heterogeneous mobility data sources facilities robust urban transportation models. Despite these attempts, however, mobility data fusion is still scarce in the literature, especially scarce in studies that tackle public health emergencies by means of mobility monitoring. Global crises, such as the COVID-19 pandemic we are facing, uniquely highlights the need for monitoring mobility dynamics in a comprehensive manner, as integrated human dynamics from multiple sources is expected to better reflect the multifaceted nature of human mobility, leading to the acquisition of overall knowledge in mobility dynamics that cover a broader spectrum of the population. At the same time, however, we argue that the representativeness of each source largely depends on the demographics of the service users in relation to the demographics of the local population. Thus, a proper weighting scheme based on their representativeness needs to be considered when multi-source mobility data are fused.

### 5.2 The luxury nature of social distancing

Our exploration of the correlation between county-level *RI* and county median income indicates that counties with higher income tend to be more responsive in terms of mobility reduction compared to the pre-pandemic baseline, evidenced by the statistically significant positive correlation between *RI* and income from all four mobility datasets. Most states show a positive difference in *RI* between their upper-income and lower-income counties, where diverging patterns in time series of mobility changes percentages can also be found. However, we need to acknowledge that correlation does not necessarily imply causation. The reasons behind the disparity in mobility patterns for upper-income and lower-income counties are multifaceted. Geographically, high-income counties in the U.S. often coexist with large cities or urban fabrics, in which the dense population is hit first and hardest by the COVID-19 pandemic. A recent study found that 54% of urban residents in the states view the disease as a major threat to day-to-day life, which compares with 42% of those living in the suburbs and just 27% of rural residents in the same states (Jones, 2020). The urban-rural discrepancy in threat awareness presumably translates to the different mobility patterns in upper-income and lower-income counties. In addition, upper-income counties within a certain state (usually with high urbanization) tend to be the job centers that receive the commuting inflow of workers from nearby lower-income counties (McKenzie, 2013). This difference in the commuting pattern is also responsible for the county-level disparity in mobility dynamics. Other possible explanations of less mobility responsiveness for lower-income counties can be traced to the policies that sometimes unintentionally create inequity among different groups (Lou et al., 2020). People respond to the mobility-restrict measures differently, largely depending on their financial resources. The outcome of these measures may reflect the preferences of the affluent but not the interests of the lower-income group (Lou et al., 2020). Given the close relationship between human mobility and the transmission of SARS-COV-2 reported from numerous studies (Gatto et al., 2020; Sirkeci and Yucesahin, 2020), the low mobility responsiveness in lower-income counties deserves more attention, as slow reduction and fast recovery in mobility may foster the second ware of infections, which could be exacerbated by the vulnerability in the low-income populations.

### 5.3 Future directions

The findings from this study point to some potential areas for future research. First, future research should investigate the representativeness of different mobility data sources, setting up the foundations for integrated multi-source mobility that reveals the travel behaviors for a broad spectrum of the population. Similarly, mobility monitoring studies need to work towards a comprehensive mobility index preparing for future emergency events, especially for global crises like the COVID-19 pandemic. Studies based on single-source data should acknowledge the underlying biases in the sample and the potential issues following these biases. Second, our study compares four open-sourced datasets that include mobility records from Descartes Labs, Apple, Google, and Twitter. Future studies can examine the similarity and dissimilarity in mobility patterns derived from other passive/active citizen-sensor data, e.g., smart cards, Wi-Fi, Bluetooth, etc. Public surveys and official mobility records from transportation services can serve as ground truth that these mobility data are compared against. Comparative studies that include multiple sources comparing against official records are still needed. The COVID-19 pandemic, an event with dramatic mobility changes on a large scale, provides a great opportunity for us to learn the strengths and weaknesses of each data source, laying the foundation for the potential multi-source integration. Third, given that county is the smallest geographical unit from Apple, Descartes, and Google mobility datasets, we explore the correlation between county-level income and county-level mobility-based responsiveness. However, changes in aggregated geographic units (e.g., from counties to Census tracts) might alter the conclusions due to the famous Modifiable Areal Unit Problem (MAUP) (Fotheringham and Wong, 1991). Future studies should investigate the disparity in mobility at various spatial units or scales. Although income is one of the fundamental factors in socioeconomic status, other factors may also contribute to the disparity in responsiveness revealed in this study, therefore deserve further investigation. Finally, the spatial incontinuity of the available counties in all four mobility datasets precludes a detailed spatial examination. However, we acknowledge that spatial non-stationarity (regional variation) may exist in the contribution of socioeconomic factors to the mobility dynamics. Future studies can consider spatial autocorrelation when using other spatially continuous mobility datasets.

## 6. Conclusion

This study reveals the similarity and dissimilarity of human mobility from various sources and the luxury nature of social distancing in the U.S. during the COVID-19 pandemic by highlighting the disparities in mobility dynamics from lower-income and upper-income counties. We collect and compute mobility data from four sources: 1) Apple mobility trend reports, 2) Google community mobility reports, 3) mobility data from Descartes Labs, and 4) mobility calculated via weighted distance from Twitter. We further design a Responsive Index (*RI*) based on the time series of mobility change percentages to quantify the general degree of mobility-based responsiveness at the U.S. county level.

The results reveal that the impact of the COVID-19 pandemic is well documented, as all mobility datasets show apparent deviations in mobility from the pre-pandemic baseline. We find statistically significant positive correlations in *RI* between either two data sources, revealing their general similarity, albeit with varying Pearson’s *r* coefficients. The *RIs* calculated from Google and Apple share the highest correlation (*r* = 0.60) presumably because both of their mobility records are from navigation applications in smartphones (i.e., Google Maps and Apple Maps). Despite the similarity, however, mobility from each source presents unique and even contrasting characteristics, demonstrating the multifaceted nature of human mobility. When correlation *RI* with income, we find that positive correlation between *RI* and income is significant in all mobility datasets, suggesting that counties with higher income tend to react more aggressively in terms of reducing more mobility in response to the COVID-19 pandemic. Despite the statistical significance, the strength of the correlation varies among datasets. The *RI* calculated from the Google dataset is strongly correlated with income (*r* = 0.65), but the correlation is rather weak between the *RI* from Apple and income. The disparity in mobility dynamics between lower-income and upper-income counties is well manifested from all mobility datasets. Most states present a positive difference in *RI* between their upper-income and lower-income counties, where diverging patterns in time series of mobility changes percentages can be found.

To our best knowledge, this is the first study that cross-compares mobility datasets from various sources during the COVID-19 pandemic. The findings contribute to gaining the knowledge of not only the characteristics of multi-source mobility data but also the mobility disparity in tandem with the wealth disparity, benefiting policy design for better mitigation of future epidemics and pandemics.

## Data Availability

Mobility datasets from Descartes Labs, Apple, and Google are open-sourced online. Twitter mobility dataset is available upon request.

https://www.descarteslabs.com/mobility/

https://www.google.com/covid19/mobility/

https://www.apple.com/covid19/mobility

## Acknowledgments

The authors want to thank Descartes Labs, Google, and Apple for making this study possible by open-sourcing their mobility datasets. The research is in part supported by NSF (2028791), University of South Carolina COVID-19 Internal Funding Initiative (135400-20-54176), and NIH (3R01AI127203-04S1). The funders had no role in study design, data collection and analysis, decision to publish, or preparation of this article.

## Declaration of competing interest

This statement confirms the authors have no competing interest in this research.

## Appendix A

**Figure A.**
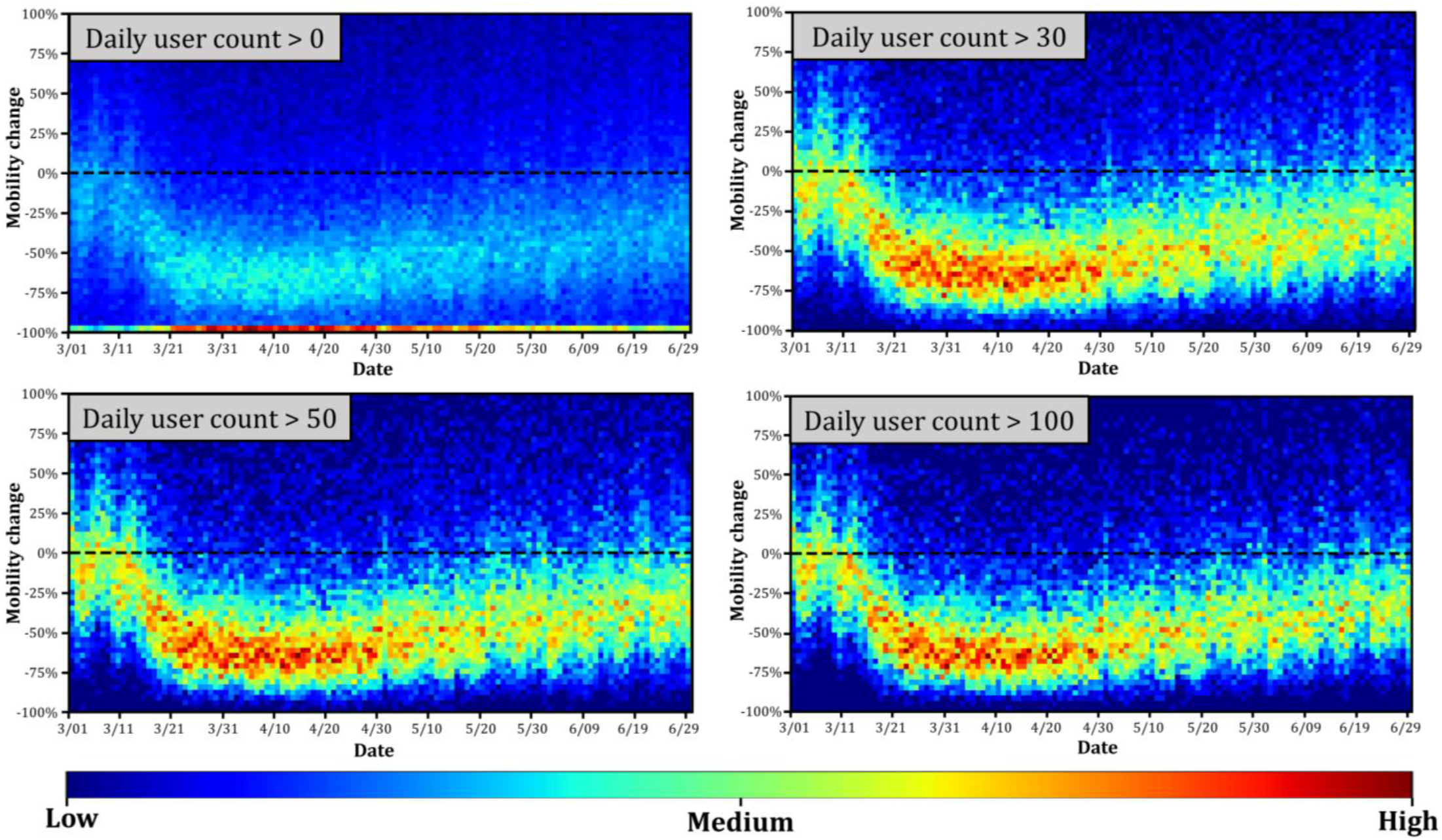
Heat map of the county-level mobility change percentages of Twitter mobility with different thresholds of daily user count (confined from -100% to 100%); (a) daily user count larger than 0; (b) daily user count larger than 30; (c) daily user count larger than 50; (d) daily user count larger than 100.

## Appendix B

**Figure B.**
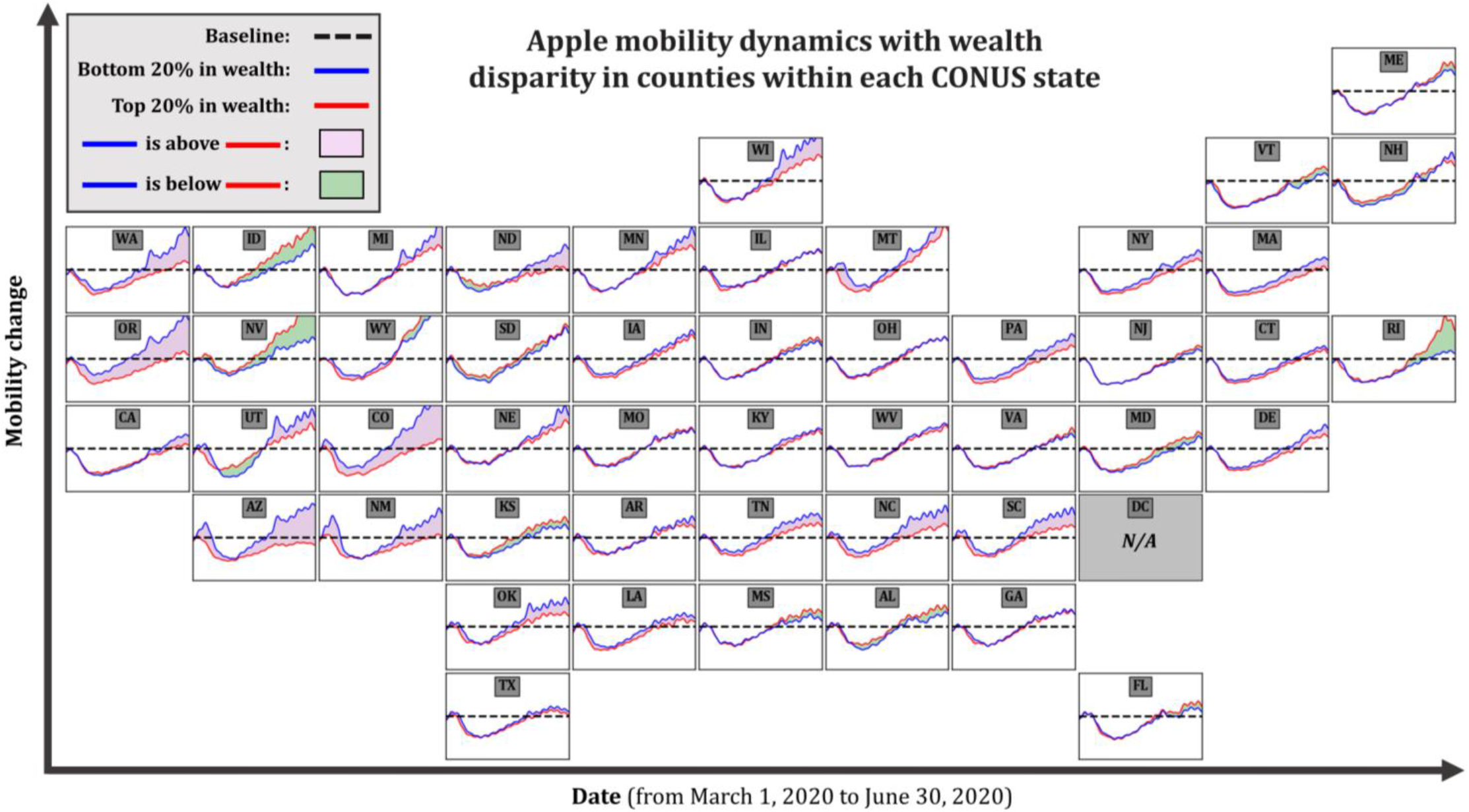
Time series of Apple mobility change percentages for the top 20% and bottom 20% counties in wealth at each CONUS state (pseudo-geographical representation). For a state with less than five available counties, counties with the most median income and the least median income are selected as the top 20% and the bottom 20%, respectively.

## Appendix C

**Figure C.**
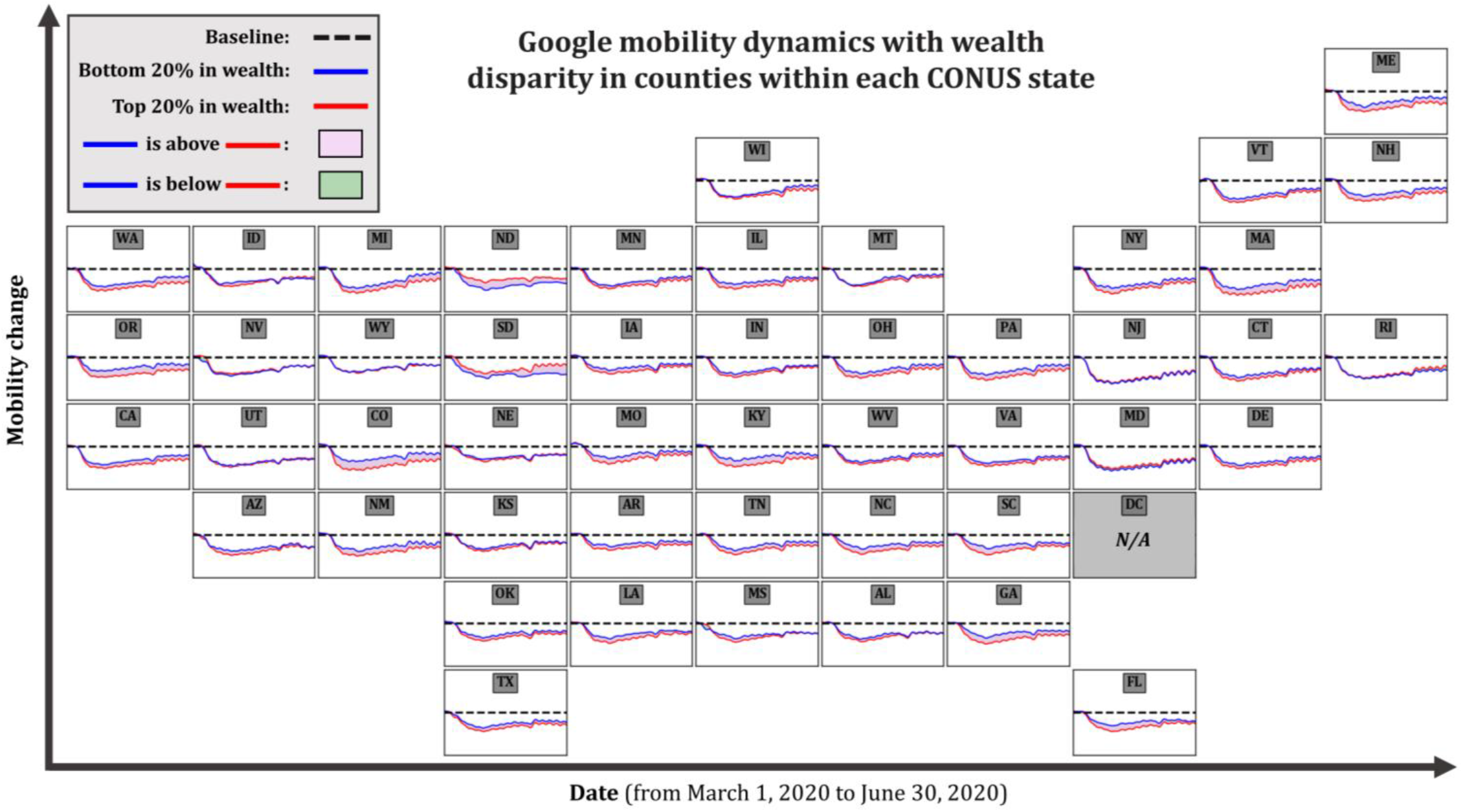
Time series of Google mobility change percentages for the top 20% and bottom 20% counties in wealth at each CONUS state (pseudo-geographical representation). For a state with less than five available counties, counties with the most median income and the least median income are selected as the top 20% and the bottom 20%, respectively.

## Notes

### Competing Interest Statement

The authors have declared no competing interest.

